# The properties of hot household hygroscopic materials and their potential use for non-medical facemask decontamination

**DOI:** 10.1101/2020.07.16.20155481

**Authors:** Marie-Line Andreola, Fréderic Becquart, Wahbi Jomaa, Paul O. Verhoeven, Gérard Baldacchino, Simon Hemour, D-Mask consortium

**Affiliations:** Université de Bordeaux, MFP, CNRS UMR 5234, 146 rue Léo Saignat, 33076 Bordeaux Cedex, France; UB’L3 TBM core, Bordeaux; Université de Lyon, CNRS, UMR 5223, Ingénierie des Matériaux Polymères, Université Jean Monnet, 42023 Saint-Etienne, France; Université de Bordeaux, CNRS UMR 5295, I2M Bordeaux, 351 cours de la Libération, 33400 Talence, France; GIMAP EA 3064 (Groupe Immunité des Muqueuses et Agents Pathogènes), University of Lyon, 42023 Saint-Etienne, France; Laboratory of Bacteriology- Virology-Hygiene, University hospital of Saint-Etienne, 42055 Saint-Etienne Cedex 02, France; Université Paris-Saclay, CEA, CNRS, Laboratoire Interactions, Dynamique et Lasers (LIDYL) - 91191 Gif-sur-Yvette, France; Université de Bordeaux, CNRS UMR 5218, Bordeaux INP, IMS, 351 cours de la Libération, 33400 Talence, France; A complete list of the D-Mask Consortium can be found in the Supporting Information

**Author notes:** Corresponding author : Simon Hemour. equally contribution authors (Wahbi Jomaa, Paul O. Verhoeven). **Author Contributions** Author contributions: M-L.A., F.B., W.J., P.V., G.B., and S.H. designed research; M-L.A., F.B., W.J., P.V., and S.H. performed research; M-L.A., F.B., W.J., P.V., G.B., and S.H. analyzed data, M-L.A.,F.B., W.J., G.B., and S.H. wrote the paper. **Competing Interest Statement:** This study has received financial support from the French State in the Frame of “Investments fof the future” program. It is partly funded by University of Bordeaux Idex OPE 2020-0208-C19 funds.

## Abstract

The wide use of facemasks through the population to prevent SARS-CoV-2 virus transmission, and its resulting mis- or even non-decontamination are challenging the management of the epidemic at a large scale. As a complement to machine-wash that wastes significant amount of water and energy, hot hygroscopic materials could be used to decontaminate non-medical facemasks in household settings. We report the inactivation of a viral load on a facial mask for an exposure of 15 minutes, with the combined effect of heat and humidity under a decaying pattern suggesting straight-forward general public deployment towards a reliable implementation by the population.

## Main Text

## Introduction

Facemasks are widely used amidst the global COVID-19 pandemic to reduce the airborne transmission in the context of social interactions (1), as high viral loads of severe acute respiratory syndrome coronavirus 2 (SARS-CoV-2) might be found in asymptomatic (2) and patients positive to the disease (3). Decontamination of used facemasks is a common practice to mitigate the shortage risk (4) and the potential ecological impact of disposable units worn by billions of daily users (1).

However, the recommended household decontamination procedures are time- and energy-intensive (5), which can potentially lead to low public acceptance. Meanwhile, it has been shown that SARS-CoV-2 can be remaining on cloth and a surgical mask for up to seven days. The infrequent decontamination that could result can then be considered as a possible indirect vector of contamination. Quick and easy decontamination methods is therefore required.

Moist heat, a combination of heat and humidity is a known treatment method of inactivating some pathogens. For example influenza viruses on stainless steel surfaces have been inactivated after being elevated during 15 minutes at the temperature of 65°C associated to a 25% relative humidity (RH) (6), while equivalent level of inactivation have been reached at 55°C and 75% RH. However, dry heat of 70°C may not be effective enough to inactivate the virus, even over a larger exposition time of an hour (7). While RH is the most reported parameter (8), absolute humidity (AH) seems to be more appropriate for predicting the influence of humidity on virus deactivation (9). During the combined heat & high absolute humidity condition, the arrangement of the lipid bilayer of the virus as well as the interactions involved in the envelope proteins could be affected (10–12). In droplets, the humidity rate and the evaporation kinetics of water can be matched (9) towards an intermediate evaporation rates and high concentration of salts, leading to virus inactivation.

Therefore, aiming at benefiting from the temperature-humidity synergy, we are reporting here the SARS-CoV-2 virus inactivation from facemask in an enclosure filled with hot hygroscopic materials available in household settings.

## Results and discussion

The following three steps procedure is tested (Figure 1-A): Two 1L polypropylene containers, each filled with 250g of hygroscopic material, are first heated in a microwave oven for 2 minutes (i). The containers are taken out of the oven, and the mask is placed in one of them, lying on the hot hygroscopic material, while the remaining material is deposited on the mask. This step has been tested with a handling time below 1:30min (ii). The container is then hermetically sealed for 15-20 min to set a high humidity atmosphere (iii).

**Figure 1.**
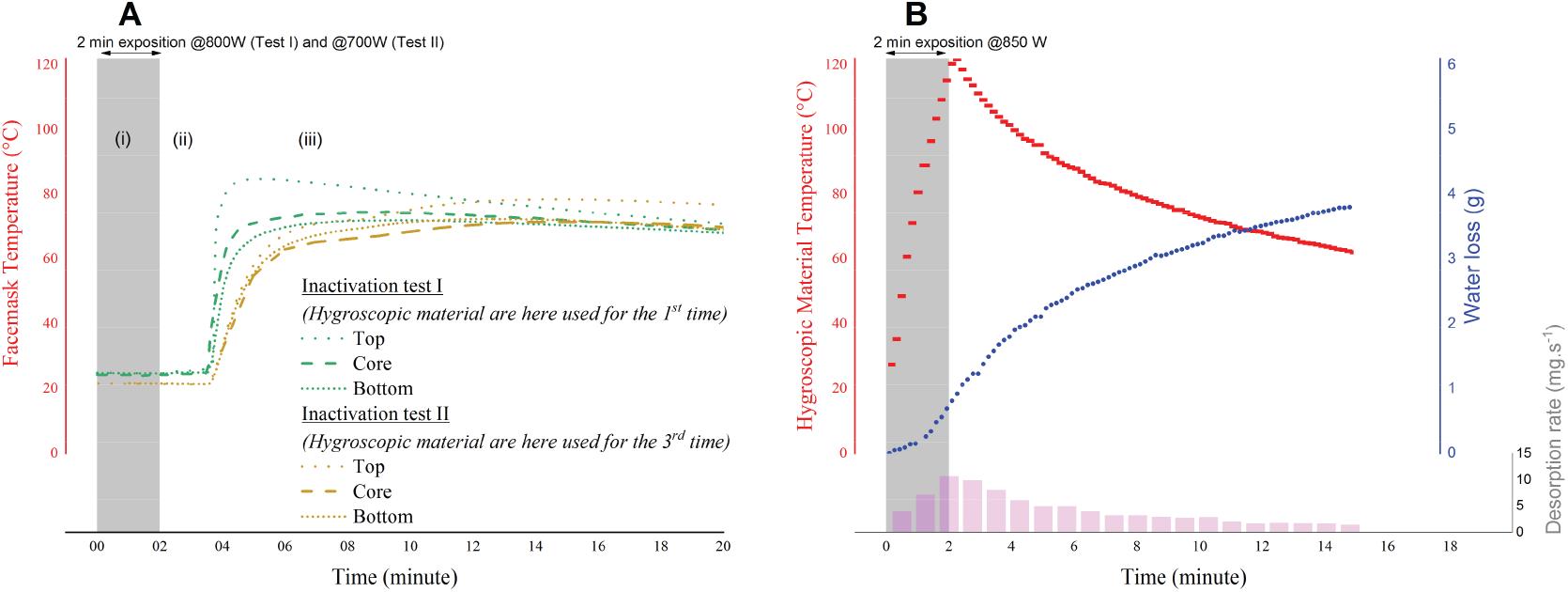
(A) Temperature evolution inside the mask under the same steaming conditions that are used for the biologic trial as pertains to Fig. 2. (B) Temperature, Water loss, and Water vapor desorption rate from initial 500g of hygroscopic material during and after a household-compatible heat and humidity decontamination process. The desorption rate is linked to the concentration of water at the surface of the medium, and thus, its pattern is a direct image of the material temperature. The fast heating process does not lead the hygroscopic medium to release all the water it contains, because the mass loss is limited by the slow material water mobility

During the first phase, the porous hygroscopic medium is exposed to micro-wave power, which results in a volumetric heat generation. Heating efficiency depends on wave interactions with polar molecules or clusters (13) described by the loss factor which is responsible for the wave attenuation and the conversion of electrical field energy into heat. In the case of dry food products, they are strongly linked to residual moisture content and starch composition (14). As a matter of fact, wheat, corn and rice starches are typically used in microwave food products formulations (15) since dielectric loss factor of these starches are commonly above 14 as compared to about 10 for water (data given at 20°C, (16)). Those materials are considered as holding potential for this decontamination method. More specifically, the process evaluated in this work is using short-cut pasta.

During the decontamination of stage (iii), the mask carrying the virus is “sandwiched” between the non-consolidated hygroscopic porous media. The relative low thickness of the mask and its low heat capacity compared to the heat capacity of the porous stack (675 J K-1 for 500 g of the chosen media, compared to 30 J K-1 for a cloth mask) ensure a fast setting of a local thermal equilibrium. The evolution of mask temperature is then governed by the evolution of porous hygroscopic media temperature. As the container is sealed during this period, heat and mass transfer with the environment are reduced leading to a slow cooling process and thus maintaining a high temperature during this phase.

Due to the hygroscopic nature of the chosen material (17), temperature elevation necessarily involves a vapor flux from the core of the material to its surface. This desorption rate ensures a high relative humidity in the vicinity of the mask during the entire protocol duration (Figure 1-B). For a typical case of using a 1L sealed box filled with 500 g pasta at 70°C, a desorption amount as low as 0.13 g is enough to generate a 100% relative humidity.

The proposed method is thus not only able to rise the temperature of the mask to a high value (Figure 1-A), but also to develop high humidity (Figure 1-B) conditions towards virus inactivation. In the hours following of the experiment, the cooled hygroscopic porous medium will smoothly reabsorbs humidity from the environment. This can be seen as a self-reactivation mechanism which allows using the material for many further decontaminations cycles. It is expected that over the decontamination cycles, heat-induced water loss from the hygroscopic medium would reduce, leading to a higher temperature. To evaluate this effect, the inactivation tests were performed with “new” material (Test I) and material used for the 3rd time (Test II). Besides, the two tests were carried out with different heating power in different BSL3 laboratories, leading to a truly two independent validations of the decontamination process.

Altogether, our results indicate that the hot hygroscopic protocol described in this work provides reliable and stable heat and humidity conditions which are efficient for inactivation of viral infectivity. This protocol holds wide potential to initiate new research in “decaying” temperature and humidity decontamination technics that are critical for the concept of frugal and reliable operation using household settings.

## Materials and Methods

For each inactivation study, 15 000 VERO E6 cells were seeded in 24 well plates for 24 hours to reach 80% confluence. A 25 µl sample of viral solution (5.13 Log TCID50/ml for test I and 4.84 Log TCID50/ml for test II) was spotted on paper patches such as antibiotic disks. Three infections were carried on in parallel (samples 1, 2 and 3) for each mask. Paper disks spotted with 25 µl of viral solution, either treated with hygroscopic materials or untreated, were used as negative control. A positive control corresponding to the spotted but untreated virus was also performed. Patches containing the virus were settled in a whole mask folds (to mimic real conditions of decontamination of an entire mask). The mask was then subject to the above described decontamination procedure. After 20 minutes for test I and 15 minutes for test II, the patches were recovered and incubated in cell culture medium for another 15 minutes, and the eluates were used to infect VERO cells. The cells were observed at 2 days, 5, 6, 7 days and 9 days (not shown) post-infection to follow the appearance of cytopathic effect. While cells with untreated virus (Fig. 2, positive control) were lysed, cells incubated with treated virus continued growing and did not show any cytopathic effect even 9 days post infection, showing the efficacy of the treatment. No toxic product was released after treatment with hygroscopic materials (negative treated control).

**Figure 2.**
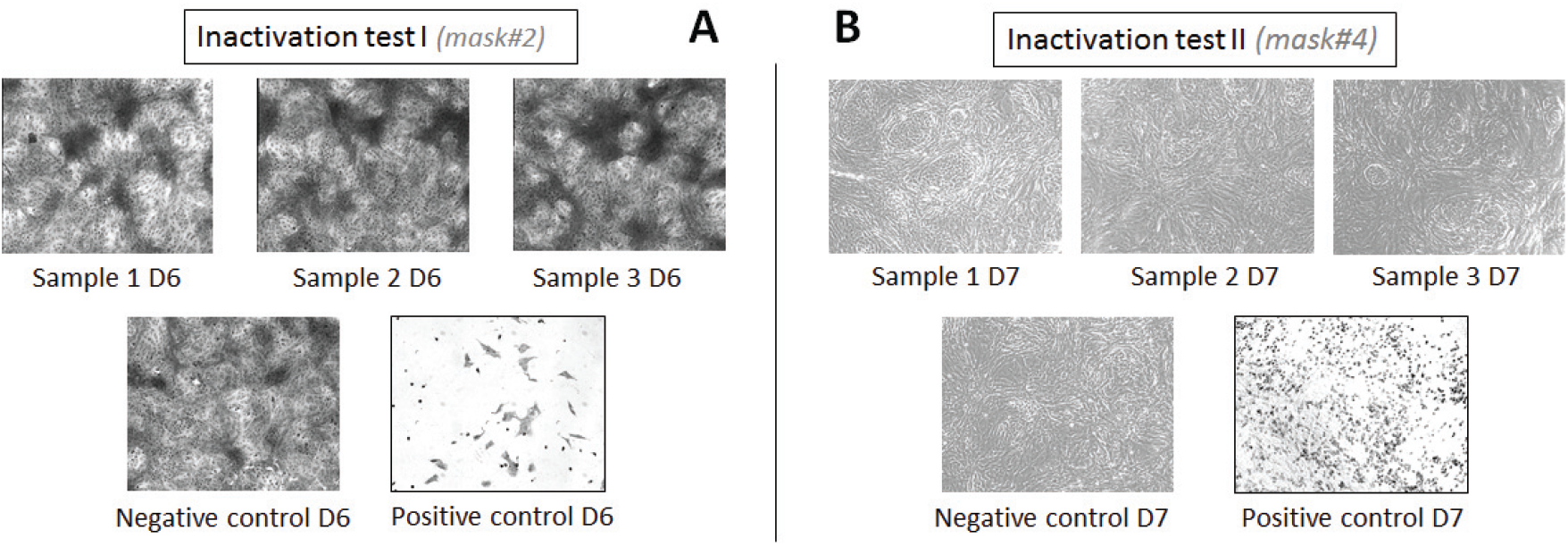
CPE at Day #6 (A), and at day #7 (B). Cells are growing in every case but for the positive control where cells are lysed. (Excerpt from the four independent inactivation tests with a total of 18 samples available in the supporting information)

RNA from inactivation test II were also extracted and amplified by qRT-PCR. Three regions of SARS CoV 2 genome were targeted (N, and two regions of Orf 1ab). While the viral RNA was detected in positive sample as expected (Ct around 15,9)), samples treated with the hygroscopic process were undetected as for the negative control and for the three amplified regions, which correspond to a loss of infectivity above 5 log.

## Data Availability

All data is included in the manuscript and supporting information

## Acknowledgments

We thank the other faculty, staff, and undergraduate research assistants, listed in the Supporting Information, who contributed many hours to support this study.

## References

1. WHO, Water, sanitation, hygiene and waste management for the COVID-19 virus. World Heal. Organ. (2020).

2. R. Wölfel, et al., Virological assessment of hospitalized patients with COVID-2019. Nature (2020) https:/doi.org/10.1038/s41586-020-2196-x.

3. J. F. W. Chan, et al., Improved molecular diagnosis of COVID-19 by the novel, highly sensitive and specific COVID-19-RdRp/Hel real-time reverse transcription-PCR assay validated in vitro and with clinical specimens. J. Clin. Microbiol. (2020) https:/doi.org/10.1128/JCM.00310-20.

4. Q. X. Ma, et al., Decontamination of face masks with steam for mask reuse in fighting the pandemic COVID-19: experimental supports. J. Med. Virol. (2020) https:/doi.org/10.1002/jmv.25921.

5. J. Yan, et al., Infectious virus in exhaled breath of symptomatic seasonal influenza cases from a college community. Proc. Natl. Acad. Sci. 115, 1081–1086 (2018).

6. J. McDevitt, S. Rudnick, M. First, J. Spengler, Role of absolute humidity in the inactivation of influenza viruses on stainless steel surfaces at elevated temperatures. Appl. Environ. Microbiol. 76, 3943–3947 (2010).

7. R. Fischer, et al., Assessment of N95 respirator decontamination and re-use for SARS-CoV-2. medRxiv, 2020.04.11.20062018 (2020).

8. K. H. Chan, et al., The Effects of Temperature and Relative Humidity on the Viability of the SARS Coronavirus. Adv. Virol. 2011, 734690 (2011).

9. K. Lin, L. C. Marr, Humidity-Dependent Decay of Viruses, but Not Bacteria, in Aerosols and Droplets Follows Disinfection Kinetics. Environ. Sci. Technol. 54, 1024–1032 (2020).

10. A. J. Prussin II, et al., Survival of the Enveloped Virus Phi6 in Droplets as a Function of Relative Humidity, Absolute Humidity, and Temperature. Appl. Environ. Microbiol. 84, e00551–18 (2018).

11. J. W. Tang, The effect of environmental parameters on the survival of airborne infectious agents. J. R. Soc. Interface 6 Suppl 6, S737–S746 (2009).

12. T. P. Weber, N. I. Stilianakis, Inactivation of influenza A viruses in the environment and modes of transmission: a critical review. J Infect 57, 361–373 (2008).

13. A. Angoy, et al., “17 - Microwave technology for food applications” in F. Chemat, E. B. T.-G. F. P. T. Vorobiev, Eds. (Academic Press, 2019), pp. 455–498.

14. X. Peng, J. Song, A. Nesbitt, R. Day, Microwave foaming of starch-based materials (I) dielectric performance. J. Cell. Plast. 49, 245–258 (2013).

15. M. K. Ndife, G. Şumnu, L. Bayindirli, Dielectric properties of six different species of starch at 2450 MHz. Food Res. Int. 31, 43–52 (1998).

16. J. Chen, et al., Development of a multi-temperature calibration method for measuring dielectric properties of food. IEEE Trans. Dielectr. Electr. Insul. 22, 626–634 (2015).

17. M. Yoshino, T. Ogawa, S. Adachi, Properties and Water Sorption Characteristics of Spaghetti Prepared Using Various Dies. J. Food Sci. 78, E520–E525 (2013).

